# Modelling the thermal inactivation of viruses from the *Coronaviridae* family in suspensions or on surfaces with various relative humidities

**DOI:** 10.1101/2020.05.26.20114025

**Authors:** Laurent Guillier, Sandra Martin-Latil, Estelle Chaix, Anne Thébault, Nicole Pavio, Sophie Le Poder, on behalf of Covid-19 Emergency Collective Expert Appraisal Group, Christophe Batéjat, Fabrice Biot, Lionel Koch, Don Schaffner, Moez Sanaa

## Abstract

Temperature and relative humidity are major factors determining virus inactivation in the environment. This article reviews inactivation data of coronaviruses on surfaces and in liquids from published studies and develops secondary models to predict coronaviruses inactivation as a function of temperature and relative humidity. A total of 102 D-values (time to obtain a log_10_ reduction of virus infectivity), including values for SARS-CoV-2, were collected from 26 published studies. The values obtained from the different coronaviruses and studies were found to be generally consistent. Five different models were fitted to the global dataset of D-values. The most appropriate model considered temperature and relative humidity. A spreadsheet predicting the inactivation of coronaviruses and the associated uncertainty is presented and can be used to predict virus inactivation for untested temperatures, time points or new coronavirus strains.

**Importance:** The prediction of the persistence of SARS-CoV-2 on fomites is essential to investigate the importance of contact transmission. This study collects available information on inactivation kinetics of coronaviruses in both solid and liquid fomites and creates a mathematical model for the impact of temperature and relative humidity on virus persistence. The predictions of the model can support more robust decision-making and could be useful in various public health contexts. Having a calculator for the natural clearance of SARS-CoV-2 depending on temperature and relative humidity could be a valuable operational tool for public authorities.

## 1. Introduction

The pandemic of coronavirus respiratory infectious disease (COVID-19) initiated in Wuhan, China in December 2019 was caused by an emergent virus named Severe Acute Respiratory Syndrome Coronavirus 2 (SARS-CoV-2). SARS-CoV-2 belongs to the order *Nidovirales*, family *Coronaviridae*. These enveloped viruses have a positive, single-stranded RNA genome (directly translated) surrounded by a nucleocapsid protein. Coronaviruses are classified into four genera: alpha (αCoV), beta (βCoV), gamma (γCoV), and delta (δCoV). SARS-CoV-2 belongs to the *Betacoronavirus* genus and the *Sarbecovirus* sub-genus.

The route of transmission of respiratory viruses is airborne via inhalation of droplets and aerosols or through contact with contaminated intermediate objects (fomites), e.g. by self-inoculation of mucous membranes (mouth, eyes) by contaminated hands (1). The transmission route for SARS-CoV-2, SARS-CoV and Middle East respiratory syndrome (MERS-CoV) is primarily airborne (2-5) while environmental contamination through surfaces is uncertain (6-8). No study has currently quantified the importance of surface contact transmission in the spread of coronavirus diseases (9). Viral genomes have been detected in the stools of COVID-19 patients and sewage (10), but the role of liquid fomites has not yet been addressed.

Working with highly virulent coronavirus requires biosafety level 3 laboratory containment conditions and since SARS-CoV2 emerged very recently, few data on its survival related to environmental conditions are available (11, 12). The use of surrogate coronaviruses has been suggested to overcome these challenges and expand the available data on coronavirus survival likelihood (13). Surrogates can be used under the assumption that they have similar physicochemical properties that mimic the viruses they represent (14, 15).

Temperature and relative humidity have been shown to impact the kinetics of inactivation of coronaviruses. Increased temperatures have been shown to increase the rate of the inactivation (11, 16), and decreased relative humidity have been associated with a reduction of coronaviruses inactivation rate on surfaces (13, 17). Inactivation rates were lower in suspensions compared to surfaces in studies that tested both suspensions and surfaces at similar temperatures (11, 18).

Hence, the prediction of the persistence of SARS-CoV-2 on fomites is essential to investigate the importance of contact transmission. This study collects available information on inactivation kinetics of coronaviruses in both solid and liquid fomites and models the impact of temperature and relative humidity on virus persistence.

## 2. Materials & methods

### 2.1. Selection of the studies

Four inclusion criteria were used to identify studies that characterized inactivation of coronaviruses according to temperature and relative humidity. Selected studies had to focus on one virus from the *Coronaviridae* family. Inactivation must have been carried out in suspensions or on inert non-porous surfaces. Only surfaces without antimicrobial properties were considered. The quantification of infectious viruses had to be assessed by cell culture, since RT-qPCR can underestimate actual virus infectivity (19, 20). Finally, the available kinetics data points should be sufficient to allow precise statistical estimation of the rate of viral inactivation without bias. In this context, kinetic data with no significant inactivation observed during the experiment or with values below the quantification limit in the first time interval were not included.

### 2.2. Data collection

The kinetics were gathered from either the figures or the tables of the selected studies. The digitize R package (21) was used to retrieve data from scatter plots in figures. This package loads a graphical file of a scatterplot (in jpeg format) in the graphical window of R and calibrates and extracts the data. Data were manually reported in R vector for data provided in tables. A key was attributed to kinetics collected in each study (Table 1). The specific list of tables and figures used for each kinetics is given in appendix 1.

**Table 1.** Characteristics of the studies that explored inactivation of infectivity of coronavirus.

| **Virus** | **Genus** | **Sub-genus** | **Strain** | **Measurement** | **Temperatures (°C)** | **Conditions associated with treatment** | **Study reference** |
| --- | --- | --- | --- | --- | --- | --- | --- |
| **BCoV** | *Betacoronavirus* | *Embecovirus* | Strain 88 | PFU in Human rectal tumor (HRT)-18 cells | 4 | Salad, MEM containing 2% FBS | (29) |
| **CCV** | *Alphacoronavirus* | *Tegacovirus* | I-71 | CRFK cells (PFU) | 60, 80<sup>a</sup> | MEM containing 2% FCS | (30) |
| **FIPV** | *Alphacoronavirus* | *Tegacovirus* | DF2-WT | Feline kidney (NLFK) cells | 54 | Basal Medium Eagle | (31) |
| **FIPV** | *Alphacoronavirus* | *Tegacovirus* | ATCC-990 | Crandell Reese feline kidney cell line | 4<sup>b</sup>, 23 | Dechlorinated, filtered tap water | (32) |
| **HCoV** | *Alphacoronavirus* | *Duvinacovirus* | 229E | Cellular infectivity in cell strain (HDCS) WI38 | 33, 37 | Maintenance medium 2% fetal calf serum | (33) |
| **HCoV** | *Alphacoronavirus* | *Duvinacovirus* | 229E | Cellular infectivity in lung cell line L132 | 21<sup>b</sup> | PBS, Earle's minimal essential medium, Earle's minimal essential medium to with added suspended cells | (34) |
| **HCoV** | *Alphacoronavirus* | *Duvinacovirus* | 229E | Cellular infectivity in lung cell line L132 | 21<sup>b</sup> | Aluminum, sponge, latex at 65% RH | (34) |
| **HCoV** | *Alphacoronavirus* | *Duvinacovirus* | 229E | CPE on MRC-5 cells | 21 | Teflon, PVC, Rubber, Steel, Plastic | (35) |
| **HCoV** | *Alphacoronavirus* | *Duvinacovirus* | 229E | - | 23 | Cell culture supernatant with or without FBS (fetal bovine serum) | (18) |
| **HCoV** | *Alphacoronavirus* | *Duvinacovirus* | 229E | MRC-5 cells (TCID-50) | 4<sup>b</sup>, 23 | Dechlorinated, filtered tap water | (32) |
| **HCoV** | *Alphacoronavirus* | *Duvinacovirus* | 229E | Cellular infectivity in lung cell line L132 | 4<sup>b</sup>, 22, 33, 37 | Earle's minimal essential medium | (36) |
| **HCoV** | *Betacoronavirus* | *Embecovirus* | OC43 | Cellular infectivity in cell strain (HDCS) WI38 | 33, 37 | Maintenance medium 2% fetal calf serum | (33) |
| **HCoV** | *Betacoronavirus* | *Embecovirus* | OC43 | Cellular infectivity in human rectal tumor cell line HRT-18 | 21<sup>b</sup> | PBS, Earle's minimal essential medium, Earle's minimal essential medium to with added suspended cells | (34) |
| **HCoV** | *Betacoronavirus* | *Embecovirus* | OC43 | Cellular infectivity in human rectal tumor cell line HRT-18 | 21<sup>b</sup> | Aluminum, sponge<sup>a</sup>, latex<sup>a</sup> at 65% RH | (34) |
| **MERS-CoV** | *Betacoronavirus* | *Sarbecovirus* | FRA2 | Cellular infectivity in Vero cells (TCID-50) | 25<sup>b</sup>, 56, 65 | Cell culture supernatant | (37) |
| **MERS-CoV** | *Betacoronavirus* | *Sarbecovirus* | HCoV-EMC/2012 | Cellular infectivity in Vero cells (TCID-50) | 20, 30 | Plastic (30%, 40% or 80% RH) | (17) |
| **MERS-CoV** | *Betacoronavirus* | *Sarbecovirus* | HCoV-EMC/2012 | Cellular infectivity in Vero cells (TCID-50) | 20, 30 | Plastic (30%, 40% or 80% RH) | (17) |
| **MHV** | *Betacoronavirus* | *Embecovirus* | - | Cellular infectivity in DBT cells | 4<sup>b</sup>, 25 | Reagent-grade water | (38) |
| **MHV** | *Betacoronavirus* | *Embecovirus* | - | Cellular infectivity in DBT cells | 4<sup>b</sup>, 25 | Lake water | (38) |
| **MHV** | *Betacoronavirus* | *Embecovirus* | - | Cellular infectivity in DBT cells | 4<sup>b</sup>, 20, 40 | Stainless steel surface with 20% humidity | (13) |
| **MHV** | *Betacoronavirus* | *Embecovirus* | - | Cellular infectivity in DBT cells | 4, 20, 40 | Stainless steel surface with 50% humidity | (13) |
| **MHV** | *Betacoronavirus* | *Embecovirus* | - | Cellular infectivity in DBT cells | 4, 20, 40 | Stainless steel surface with 80% humidity | (13) |
| **MHV** | *Betacoronavirus* | *Embecovirus* | MHV-2 | Cellular infectivity in DBT cells (PFU) | 40<sup>b</sup>, 60, 80<sup>a</sup> | MEM containing 2% FCS | (30) |
| **MHV** | *Betacoronavirus* | *Embecovirus* | MHV-N | Cellular infectivity in DBT cells (PFU) | 40<sup>b</sup>, 60, 80<sup>a</sup> | MEM containing 2% FCS | (30) |
| **MHV** | *Betacoronavirus* | *Embecovirus* | A59 | Plaque assay on L2 cells | 10<sup>b</sup>, 25 | Pasteurized wastewater | (39) |
| **PEDV** | *Alphacoronavirus* | *Pedacovirus* | V215/78 | PFU on Vero cells | 50 | Diluted medium for virus replication | (40) |
| **PEDV** | *Alphacoronavirus* | *Pedacovirus* | CV777 | Vero cells (TCID-50) | 40, 44, 48 | MEM at pH 7.2 | (41) |
| **PEDV** | *Alphacoronavirus* | *Pedacovirus* | CV777 | Vero cells (TCID-50) | 4<sup>b</sup>, 44<sup>b</sup>, 48 | Medium at pH 7.5 | (42) |
| **SARS-CoV** | *Betacoronavirus* | *Sarbecovirus* | FFM-1 | Cellular infectivity in Vero cells | 56 | Cell culture supernatant with or without FBS (fetal bovine serum) | (18) |
| **SARS-CoV** | *Betacoronavirus* | *Sarbecovirus* | Urbani | Cellular infectivity in Vero cells | 56, 65, and 75<sup>a</sup> | DMEM (Dulbecco's modified Eagle's medium) | (43) |
| **SARS-CoV** | *Betacoronavirus* | *Sarbecovirus* | HKU39849 | Cellular infectivity in FRH-K4 (TCID-50) | 28, 33, 38 | Plastic stored at 95% RH | (44) |
| **SARS-CoV** | *Betacoronavirus* | *Sarbecovirus* | HKU39849 | Cellular infectivity in FRH-K4 (TCID-50) | 28<sup>b</sup>, 33, 38 | Plastic stored at 80-89% RH | (44) |
| **SARS-CoV** | *Betacoronavirus* | *Sarbecovirus* | GVU6109 | Cellular infectivity in Vero cells (TCID-50) | 20 | VTM medium | (45) |
| **SARS-CoV** | *Betacoronavirus* | *Sarbecovirus* | GVU6109 | Cellular infectivity in Vero cells (TCID-50) | 4, 20 | Nasopharyngeal aspirate (NPA), throat and nasal swab<br>(TNS) or VTM medium | (45) |
| **SARS-CoV** | *Betacoronavirus* | *Sarbecovirus* | Tor2 AY274119.3 | Cellular infectivity in Vero cells (TCID-50) | 22 | Plastic and stainless steel stored at 40°C | (12) |
| **SARS-CoV** | *Betacoronavirus* | *Sarbecovirus* | Utah | Cellular infectivity in Vero cells (TCID-50) | 58, 68 | Iscove's 4% FCS medium | (46) |
| **SARS-CoV** | *Betacoronavirus* | *Sarbecovirus* | Utah | Cellular infectivity in Vero cells (TCID-50) | 22 | Glass surface store at 10-25% RH | (46) |
| **SARS-CoV** | *Betacoronavirus* | *Sarbecovirus* | Hanoi | Cellular infectivity in Vero cells (TCID-50) | 56 | Minimum essential medium | (47) |
| **SARS-CoV2** | *Betacoronavirus* | *Sarbecovirus* | - | Cellular infectivity in Vero cells (TCID-50) | 4, 22, 37, 56, 70<sup>a</sup> | Virus transport medium | (11) |
| **SARS-CoV2** | *Betacoronavirus* | *Sarbecovirus* | - | Cellular infectivity in Vero cells (TCID-50) | 22 | Plastic and stainless steel at 65% HR | (11) |
| **SARS-CoV2** | *Betacoronavirus* | *Sarbecovirus* | WA1-2020 (MN985325.1) | Cellular infectivity in Vero cells (TCID-50) | 22 | Plastic and stainless steel stored at 40°C | (12) |
| **SARS-CoV2** | *Betacoronavirus* | *Sarbecovirus* | - | Cellular infectivity in Vero cells (TCID-50) | 56, 65<sup>a</sup> | Cell culture supernatants | (48) |
| **SARS-CoV2** | *Betacoronavirus* | *Sarbecovirus* | - | Cellular infectivity in Vero cells (TCID-50) | 65, 95<sup>a</sup> | Nasopharyngeal samples | (48) |
| **SARS-CoV2** | *Betacoronavirus* | *Sarbecovirus* | - | Cellular infectivity in Vero cells (TCID-50) | 56 | Sera | (48) |
| **TGEV** | *Alphacoronavirus* | *Tegacovirus* | D52 | Cellular infectivity in RPTg cells | 31, 35, 39, 43, 47, 51 and 55 | In HEPES solution at pH 7 | (16) |
| **TGEV** | *Alphacoronavirus* | *Tegacovirus* | D52 | Cellular infectivity in RPTg cells | 35, 39, 43, 47, and 51 | In HEPES solution at pH 8 | (16) |
| **TGEV** | *Alphacoronavirus* | *Tegacovirus* | - | Cellular infectivity in ST cells | 4<sup>b</sup>, 20, 40 | Stainless steel surface with 20% humidity | (13) |
| **TGEV** | *Alphacoronavirus* | *Tegacovirus* | - | Cellular infectivity in ST cells | 4, 20, 40 | Stainless steel surface with 50% humidity | (13) |
| **TGEV** | *Alphacoronavirus* | *Tegacovirus.* | - | Cellular infectivity in ST cells | 4, 20, 40 | Stainless steel surface with 80% humidity | (13) |
| **TGEV** | *Alphacoronavirus* | *Tegacovirus.* | - | Cellular infectivity in ST cells | 4<sup>b</sup>, 25 | Reagent-grade water | (38) |
| **TGEV** | *Alphacoronavirus* | *Tegacovirus.* | - | Cellular infectivity in ST cells | 4<sup>b</sup>, 25 | Lake water | (38) |

<sup>a</sup> not included: limit of quantification reached for the first sample time
<sup>b</sup> not included: not enough decrease observed during experimentation
- data not specified

### 2.3. Modelling of inactivation

A simple primary model was used for describing each inactivation kinetics. The D-values (or decimal reduction times) were determined from the kinetics of the log_10_ number of infectious viruses (*N*) over time at each experimental temperature. *D* is the inverse of the slope of the inactivation kinetics.

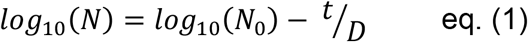

Several secondary models describing the impact of temperature (T) and relative humidity (RH) on *D* values were tested. The gamma concept of inactivation was used (22, 23). In this approach, the inactivation of a microbial population could be estimated by:

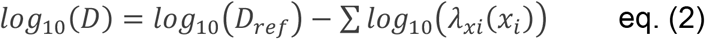

Where *λ_xi_* quantifies the influence of each environmental factors (*X_i_* corresponds to temperature and relative humidity in this study) on the microbial resistance (*D*_ref_) observed in reference conditions.

Based on eq. (2), five different secondary models were established.

Model #1 is the classical Bigelow model (24). It models only the effect of temperature. The z_T_, the increase of temperature which leads to a tenfold reduction of *D*, value was determined as the negative inverse slope of the plot of log_10_(*D*) versus temperature. z_T_ is the increase of temperature which leads to a ten-fold reduction of the decimal reduction time. *T*_ref_ is the reference temperature (set to 4°C in our study) and log_10_(*D*_ref_) is the log_10_(*D*) at *T*_ref_.

Model #1

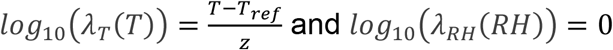

Model #2 considers the effect of temperature, however *D* values were fitted according to temperature using a semi-log approach, derived from Mafart *et al*. (2001).

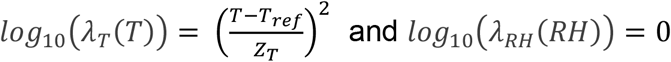

Model #3 is similar to model #2 but the shape parameter n was estimated instead of being set to 2.

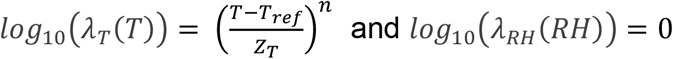

The last two models (#4 and #5) include the effect of relative humidity together with the effect of temperature. In model #4, the shape parameter for temperature was set to 2.

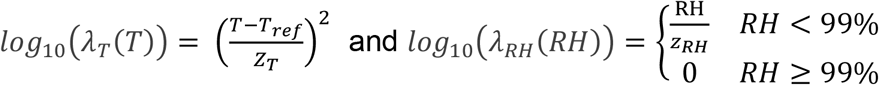

In model #5, n is a model parameter to be estimated.

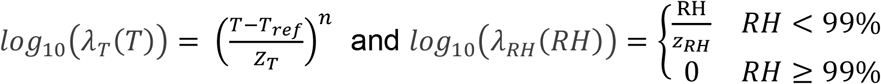

In models #4 and #5, z_RH_ is the increase of relative humidity, which leads to a ten-fold reduction of the decimal reduction time.

### 2.4. Model’s parameters estimation

The model’s parameters were fitted with nls() R function. Confidence intervals of fitted parameters were assessed by bootstrap using nlsBoot() function from nlsMicrobio R package (25). The five models were compared according to penalized-likelihood criteria, the Aikaike information criterion (AIC) (26) and Bayesian information criterion (BIC) (27).

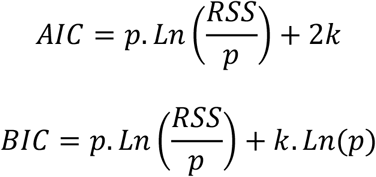

Where RSS is the residual sum of squares, *p* is the number of experimental points and *k* the number of parameters in the model. The lower the AIC and BIC, the better the model fits the dataset.

### 2.5. Data availability

The detailed information on the tables and figures where the data were collected are given in appendix 1. All the scripts and data used to prepare figures and tables of this manuscript are available in a Github repository (28).

## 3. Results

### 3.1. Literature review results

Table 1 shows the detailed characteristics of the twenty-six studies that characterized inactivation of a virus from the *Coronaviridae* family according to temperature and or relative humidity. Some kinetics were not appropriate for characterizing inactivation rate either because the duration of the experiments was too short to observe any significant decrease of virus infectivity, or because the quantification limit was reached before the first time point (Table 1). A total of 102 estimates of D-value were collected from 25 of the 26 studies (Appendix 1). These kinetic values represent 605 individual data points. For each curve, a D-value (i.e. decimal reduction time) was estimated. The 102 D-values are given in Appendix 1. Among the 102 kinetic values, 44 are from members of the *Alphacoronavirus* genus including one from Canine coronavirus (CCV), two for the feline infectious peritonitis virus (FIPV), five for the porcine epidemic diarrhea virus (PEDV), 14 for the Human coronavirus 229E (HCoV-229E) and 22 from the porcine transmissible gastroenteritis coronavirus (TGEV). The remaining 58 kinetics are related to the *Betacoronavirus* genus, including two Human coronavirus - OC43 (HCoV-OC43), two for the bovine coronavirus, 13 for the murine hepatitis virus (MHV), eight for the MERS-CoV, 22 for the SARS-CoV and 11 for the SARS-CoV-2.

Figure 1 shows the 102 estimates of D-values, including 40 values on inert surfaces and 62 values in suspension from temperatures ranging from 4°C to 68°C. Different suspensions were noted, but most were laboratory media (Table 1).

**Figure 1.**
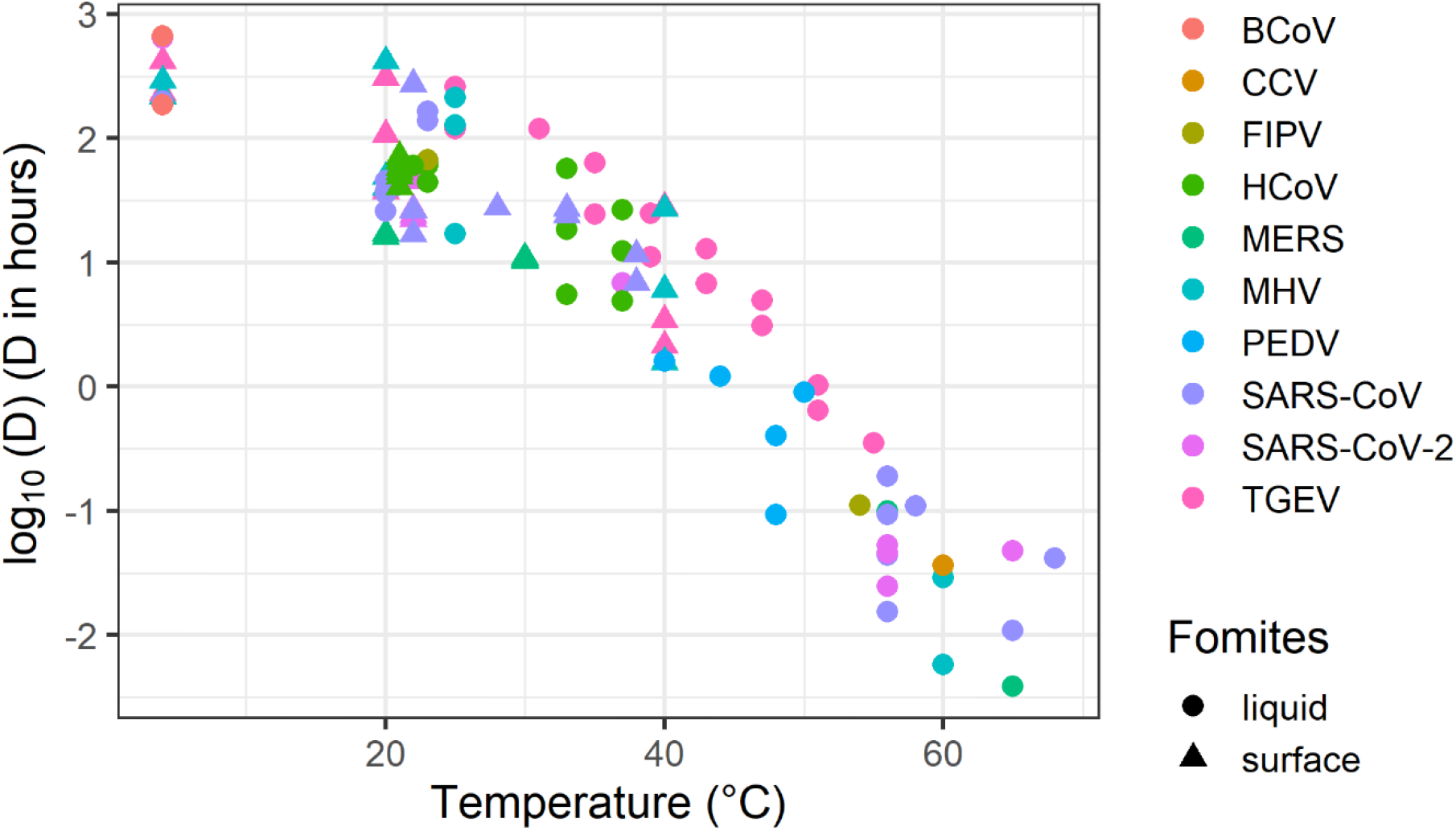
Decimal reduction times of ten coronaviruses according to temperature in suspension or on inert surfaces.

### 3.2. Modelling the inactivation

The 102 D-values were fitted with five different models. Table 2 shows the performance of these models to describe D-values according to temperature and relative humidity. For the tested range of temperatures (between 4 and 68°C), model #1 (the classical Bigelow model) based on a log-linear relation between D-values and temperature does not perform as well as model #2 that considers a linear second-degree equation. Model #3 offers a further refinement over model #2 by also fitting the degree of the equation (n parameter). The fitted value of *n* was equal to 1.9 with a confidence interval that includes 2 *(i.e*. model #2). Accordingly, the values taken by the parsimony criterions for model selection AIC and BIC for model #2 and #3, indicate that *n* can be set to 2.

**Table 2.** Characteristics of the different models fitted to the 102 decimal reduction time data of coronaviruses according to temperature (TVef set at 4°C) and relative humidity.

| Model | Fitted parameters | Best fit values<br>[95%CI bootstrap intervals] | BIC | AIC |
| --- | --- | --- | --- | --- |
| Model #1 | log10Dref<br>$z_T$ | 3.1 [2.8 - 3.3]<br>13.8 [12.7 - 15.1] | -124.7 | -130.0 |
| Model #2 | log10Dref<br>$z_T$ | 2.2 [2.1 - 2.3]<br>29.4 [28.4 - 30.5] | -160.6 | -165.9 |
| Model #3 | log10Dref<br>$z_T$<br>$n$ | 2.3 [2.1 - 2.6]<br>27.7 [23.2 - 31.6]<br>1.9 [1.5 - 2.2] | -156.7 | -164.6 |
| Model #4 | log10Dref<br>$z_T$<br>$z_{RH}$ | 2.3 [2.2 - 2.5]<br>29.1 [28.1 - 30.1]<br>341.4.7 [190.1 - 5631.4] | -160.2 | -168.0 |
| Model #5 | log10Dref<br>$z_T$<br>$z_{RH}$<br>$n$ | 2.4 [2.2 - 2.6]<br>27.5 [23.6 - 31.2]<br>330.7 [182.8 - 7020,1]<br>1.9 [1.6 - 2.2] | -156.2 | -166.6 |

Figure 2 illustrates the performance of models #1 (Fig. 2A), #2 (Fig. 2B) and #3 (Fig. 3C) for which only temperature effect is considered for predicting D-values.

**Figure 2.**
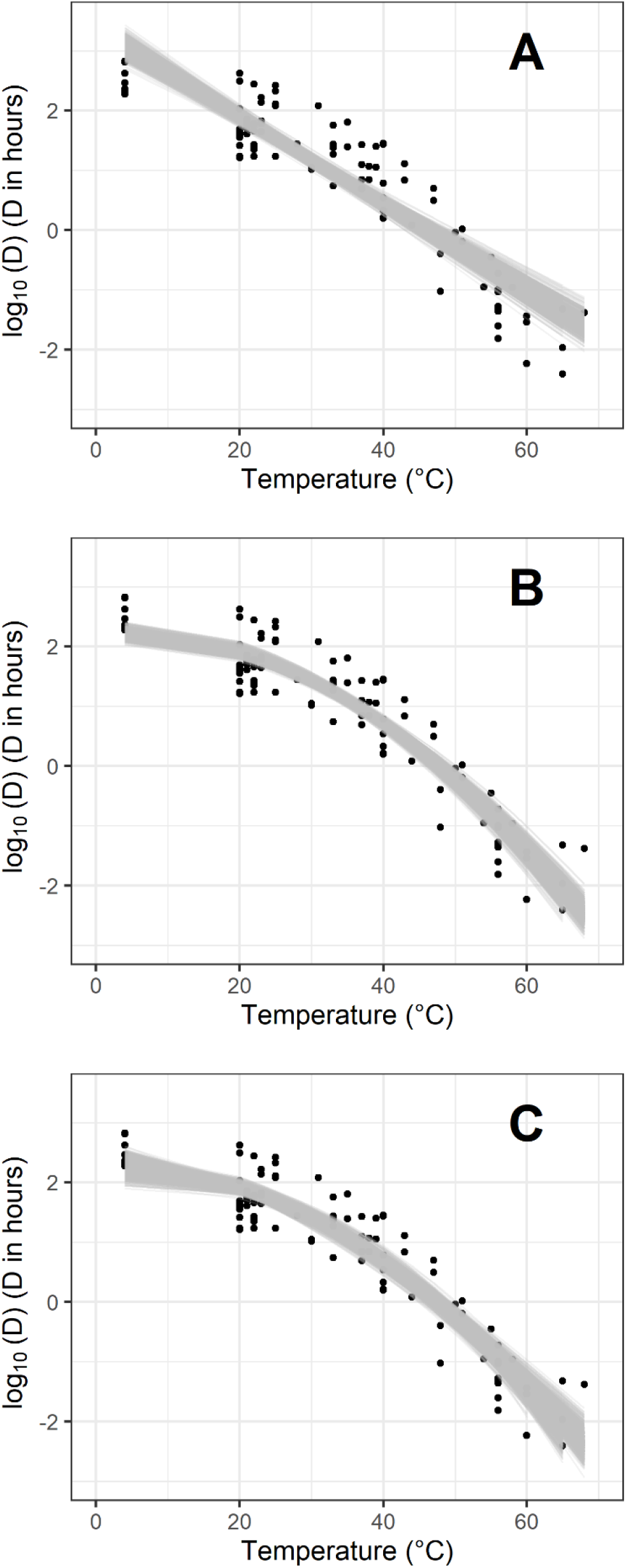
Observed (points) and fitted (grey lines) log decimal reduction time values according to temperature for model #1 (A), model #2 (B) and #3 (C). One thousand (1000) bootstrap values of uncertainty characterisation are shown. Estimates of model parameters are given in Table 2.

Table 2 demonstrates that the inclusion of relative humidity should be considered. Models #4 and #5 that describe the D-values according to temperature and relative humidity were more appropriate models than models #1, #2 and #3 with a decrease of AIC of more than 2 points in comparison with other models (49). The estimated value for the shape parameter in Model #5 is not different from the value two. According to BIC criterion, models #4 and model #2 were the most appropriate. Appendix 2 shows a further comparison of these two models. According to the two parsimony criterions, the best model overall was Model #4. Figure 3A shows the prediction of inactivation rate according to T and RH for this model.

**Figure 3.**
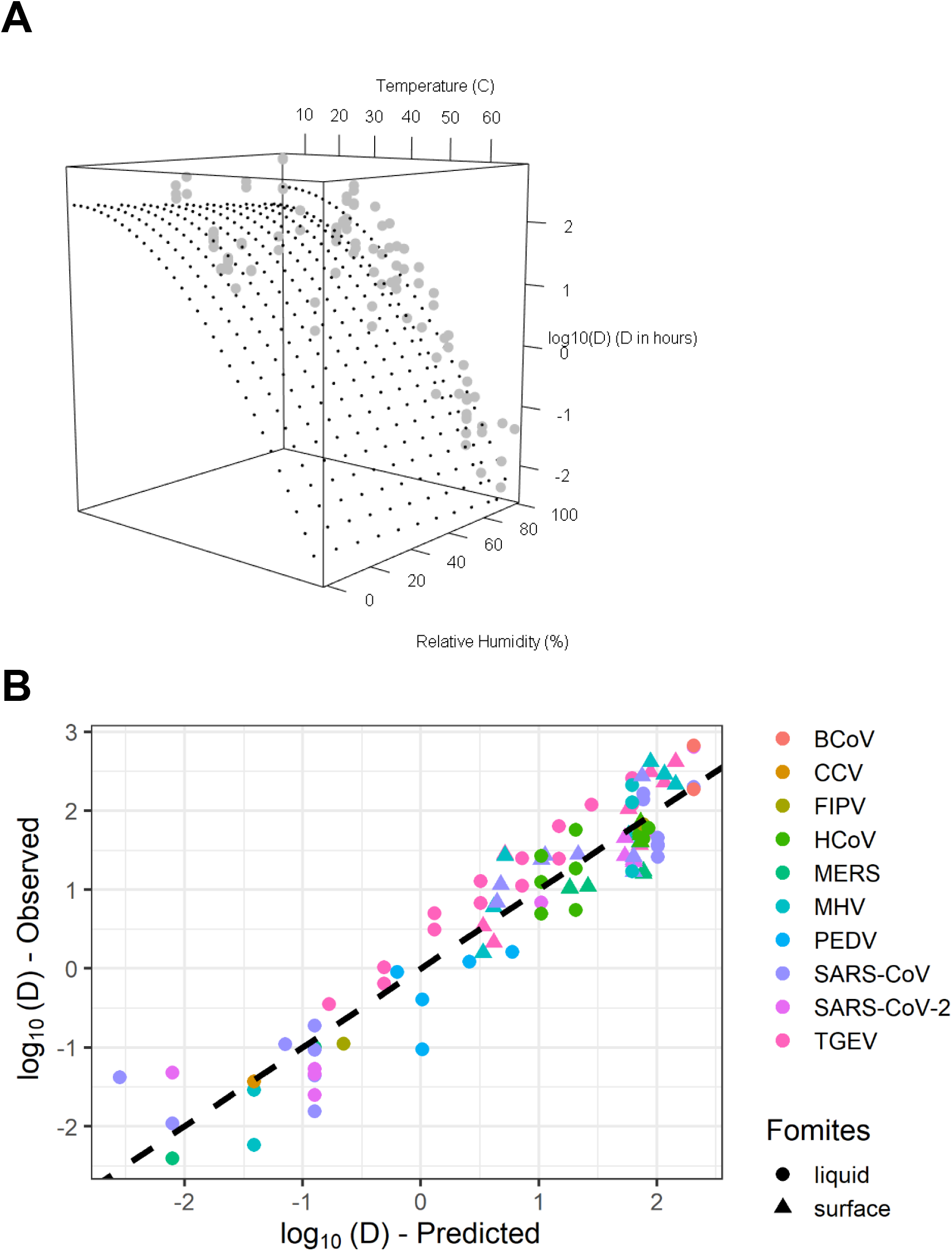
(A) Observed inactivation rate values (grey points) according to temperature and relative humidity and Model #4 predictions (dashed black points). (B) Scatter points of observed versus predicted D-values. The dashed line represents a perfect match between observations and predictions.

### 3.3. Potential use of the model

An Excel spreadsheet implementing model #4 has been prepared and is available in Appendix 3. The spreadsheet can be used to estimate the number of decimal reductions of infectivity of coronaviruses according to user defined time, temperature and relative humidity. For example, the predicted inactivation for a temperature of 70°C for 1 minute in liquid is −11.8 log10 with a 95% CI [−6.4; −22.1]. The spreadsheet also allows an estimate of the time necessary to reach a target number of decimal reductions of infectivity with a certain confidence level. For example, the time to reach a 5 log10 inactivation at 20°C and 20% of relative humidity is 438 h with a 95% CI of [339; 569].

## 4. Discussion

Our study identified 102 kinetic values for inactivation of coronaviruses on surfaces and in suspensions. The included studies cover those identified in three recently published articles that conducted a systematic review on coronaviruses inactivation (50-52). These data were used to suggest a novel inactivation model specific to the *Coronaviridae* family. The modelling approach identified temperature and relative humidity as major factors needed to predict infectious coronavirus persistence on fomites.

The log_10_ of D values was not linearly related to temperature in the range of temperatures studied (4 - 68°C). Bertrand et al. (15) made a similar observation in a meta-analysis for virus and phage inactivation in foods and water and proposed two different models on either side of the threshold temperature of 50°C. Laude (16) suggested a similar approach for TGEV with a threshold temperature at 45°C (16). The modelling approach we used in our study allows fitting the inactivation values with a single relation. In another meta-analysis on inactivation of viruses, Boehm et al. (20) did not observe such different trends but also studied a smaller temperature range (4 - 29°C). In the highest range of temperature (above 60°C), coronaviruses are found to be far less heat resistant than non-enveloped viruses (53).

The present modelling approach considers the non-monotonous impact of relative humidity on inactivation. Coronaviruses persisted better at low RHs and at 100% RH, than for intermediate RHs. Lin and Marr (54) recently observed the same relation for two bacteriophages, where the observed RH where survival was worst is close to 80% while in the present study, the less favourable condition for coronaviruses was set to 99%. Another study has confirmed that low RH makes viruses more resistant to thermal inactivation (55).

As noted in the methods, all the kinetic values analyzed were established based on the quantification of coronavirus infectivity with cell cultures. The model prediction did not include other inactivation results from methods combining dyes with RT-qPCR. This method (although more appropriate than classical RT-qPCR) can underestimate virus infectivity (19, 20).

Our findings suggest that persistence potential of different coronaviruses is similar. It confirms previous finding that advocates for the use of surrogates’ coronavirus such as TGEV (38). This could considerably simplify the acquisition of relevant data for persistence potential for other environmental factors. The data analyzed here only include *Alpha-* and *Betacoronavirus*, as no data for the two other major sub-genus, *Delta-* and *Gammacoronavirus*, were identified. Inclusion of such data would help to challenge the present model robustness.

The models developed in our study are specific to viruses from the *Coronaviridae* family. Several studies on the inactivation of other viruses have suggested that the impact of temperature can be modelled, as a whole, with a unique parameter (15, 20, 56). Variability of behavior by virus type has been observed and model parameters to account these differences have been proposed (20, 56), e.g. non-enveloped viruses are known to show greater persistence in the environment (56). Like a recently proposed model for SARS-CoV-2 (57), our model takes into consideration of relative humidity in the prediction of inactivation. This integration is of high interest in the perspective of assessment of seasonality on virus persistence (58).

It’s also worth noting our model is specific to fomites. Survival kinetics in fecal materials were identified (45) but not considered for inclusion. The level of matrix contamination with fecal materials has been shown to significantly increase the inactivation rate of viruses (56), so by excluding these data, model predictions are biased to be fail-safe. Inactivation data on porous surfaces were also not considered since it may be difficult to determine if any measured inactivation is associated with real loss of infectivity or difficulty in recovering viruses absorbed inside the porous material. That said, there is no reason to consider that model predictions for coronaviruses are not pertinent to survival on porous material (e.g. face masks).

Inactivation on anti-microbial surfaces, such as copper and silver, was also not considered. For the same reason, model predictions are fail safe as surfaces including copper or other antimicrobial compounds increase the inactivation rate of coronaviruses (12, 35).

The predictions of the present model could support more robust decision-making and could be useful in various contexts such as blood safety assessment (59) or validation of thermal inactivating treatments for room air, surfaces or suspensions. Indeed, an important issue is the possibility of reusing privates or public offices, rooms of hotels, or vehicles that are difficult to decontaminate. Moreover, many devices like electronics or more sensitive materials, are not suitable for chemical decontamination processes which could make them inoperative. Another aspect of decontamination is the economical challenge as large scale decontamination of buildings can cost billions of dollars (60). Furthermore, the use of detergents and/or disinfectants may have environmental consequences. Thus the large scale decontamination of surfaces for SARS-CoV-2, that are not necessarily in contact with people may not be required. For these reasons the waiting time needed before handling suspected contaminated materials in absence of decontamination is more than ever an important question. Having a calculator for the natural clearance of SARS-CoV-2 depending on temperature could be a valuable operational tool for public authorities (57).

The present model also opens the way for risk assessment for SARS-CoV-2 transmission through contact (61). Further model developments including data on matrix pH, salinity and exposure to visible and UV light would also be important to consider (56).

## Data Availability

The datasets analysed during the current study are available in the https://github.com/lguillier/Persistence-Coronavirus repository

## 5. Acknowledgements

The Covid-19 Emergency Collective Expert Appraisal Group members included the co-authors LG, S.M-L, E.C, N.P, S.L.P, M.S. and (in alphabetical order): Paul Brown, Charlotte Dunoyer, Florence Etore, Elissa Khamisse, Meriadeg Legouil, François Meurens, Gilles Meyer, Elodie Monchatre-Leroy, Gaëlle Simon and Astrid Vabret.

